# Personality traits and other factors associated with psychotropic medication non-adherence at two hospitals in Uganda. A cross-sectional study

**DOI:** 10.1101/2024.04.05.24305308

**Authors:** Emmanuel Niyokwizera, David Nitunga, Joshua Muhumuza, Raissa Niyubahwe, Nnaemka Chukwudum Abamara, Joseph Kirabira

## Abstract

**Background:** Mental illnesses, like other chronic illnesses, require medications for both immediate, short term and long term treatment. Medication adherence is the first and most important factor for better treatment outcome. Non-adherence to psychotropic medications is associated with relapse, readmission, and early death. The beliefs about medication which influence non-adherence to medications are moderated by specific personality traits. Sociodemographic and clinical factors can also influence non-adherence psychotropic medications. Non-adherence to psychotropic medications is high in Africa but there is paucity of published studies on the level of psychotropic medication non-adherence and associated personality traits in Uganda.

**Aim:** To determine the prevalence of psychotropic medication non-adherence and associated personality traits among people with mental illness attending Kampala International University Teaching Hospital (KIU-TH) and Jinja Regional Referral Hospital (JRRH).

**Methods:** This study employed a hospital-based cross-sectional design. 396 adult patients suffering from mental illness were collected from KIU-TH and JRRH outpatient clinics. Medication adherence was assessed using Medication Adherence Rating Scale (MARS) while personality traits were assessed by the short form of Big Five Inventory (Ten Items Personality Inventory). We first assessed sociodemographic and clinical factors influencing psychotropic medication non-adherence in our study (confounders). A questionnaire with sociodemographic and clinical information was also used. Logistic regression was used to assess personality traits and other factors associated with psychotropic medication non-adherence.

**Results:** The majority of the study participants were males (59.1%), from rural areas (74.2%), with secondary educational level (47.5%) and unemployed (44.9%). The prevalence of psychotropic medication was 46.21%. Poor family support (aOR= 6.915, CI=3.679-12.998, P<0.001), belief in witchcraft/sorcery (aOR=2.959, CI=1.488-5.884, P=0.002), experiencing side effects (aOR=2.257, CI=1.326-3.843, P=0.003), and substance use (aOR=4.174, CI=2.121-8.214, P<0.001) were factors significantly associated with psychotropic medication non-adherence. The personality traits significantly associated with psychotropic medication non-adherence after controlling for the confounders were neuroticism (aOR=7.424, CI=3.890-14.168, P<0.001) and agreeableness (aOR=0.062, CI=0.024-0.160, P<0.001).

**Conclusion:** Medication non-adherence was high. Non-adherent patients were more likely to have predominant neuroticism personality traits. Medication non-adherence was shown to be less common in individuals with agreeableness personality traits. Other factors associated with psychotropic medication non-adherence were poor social support, witchcraft beliefs, presence of side effects and substance use. Reinforced psycho-education should be given to patients with high risk of being non-adherent to psychotropic medications.

## Background

Worldwide, psychiatric disorders are among the common public health problems and are responsible for more economic burden than somatic disease like diabetes and cancer (1). In Sub-Saharan African, by 2050, the burden due to psychiatric disorders and substances use disorders is estimated to increase globally by 130% and specifically by 139% in Eastern Africa if there is no change in the prevalence and management of these disorders (2). The 2017 report of WHO indicated that globally, 80% of people with mental disorders are living in low- and middle-income countries (LMIC) which include Uganda and up to 75% of affected persons in many LMIC did not have access to the needed treatment (3). In Uganda, the prevalence of mental disorders is 22, 9% in children and 24, 2% in adults and around 80% of population with psychiatric disorders used both traditional and modern treatment which can predict a poor adherence to medications (4).

Patients with psychiatric disorders are more likely to be non-adherent to treatment than those with medical diseases (5). Worldwide, the prevalence of non-adherence to medication among psychiatric disorders is estimated at 49% and factors associated with non-adherence varies in different countries(5). In Africa the prevalence of adherence is 48% but varies between 20-80 %(5–7). Medication non-adherence is a complex and multifaceted health-care issue and is influenced by different factors divided in patient-related factors, physician-related factors, health system/team building related factors (8)

Sociodemographic factors (age, gender, marital status, occupation, family and social support, etc.), substance abuse, beliefs about mental illness are important factors associated with non-adherence related to the patient (9). In Uganda, traditional beliefs about mental disorders are still present in society and could influence medical treatment adherence(10). Illness related factors like side effects of medications, beliefs about the illness, treatments complexity and lack of insight are barriers to medication adherence(11). The beliefs about medication which influence non adherence are moderated by specific personality traits (12). Personality trait can predict healthcare utilization as well as health outcomes (13).In Italy neuroticism and conscientiousness were the most personality traits associated with psychotropic medication non adherence. High levels of conscientiousness traits were related to poorer understanding of the clinical information received from healthcare professionals (14). In India, conscientiousness and agreeableness personality traits were associated with medication adherence (15).

Non-adherence to psychiatric drugs is associated with relapse, re-hospitalization and premature death (11). Despite the free access to medications in some regions, non-adherence to psychotropic medications is still high in Eastern Africa but there is currently paucity of published studies on the level of psychotropic medication non-adherence and associated factors in Uganda (16).

Assessment of medication adherence could help to understand the reasons for non-compliance in people with mental disorders and create the framework for therapies aimed at boosting adherence. Therefore, this study aimed at determining the prevalence of psychotropic medication non-adherence and associated personality traits (as well as other factors) in Uganda.

## 2. Methods

### 2.1. Aim, design, setting and study population

Our aim was to determine the prevalence of psychotropic medication non-adherence and associated personality traits among patients with mental illness. Before reaching that objective, we first assessed others possible factors (confounders) that could influence medication non-adherence in our study participants. A descriptive cross-sectional study employing quantitative method was conducted from 1^st^ July to 1^st^ September 2023 among people with mental disorders attending psychiatric outpatient clinics at Kampala International University-Teaching Hospital and Jinja Regional Referral Hospital. Patients diagnosed with mental disorder and taking psychotropic medications for at least 6 months, aged 18 years old and above were included in the study. Patient with active psychiatric symptoms were excluded from the study.

### 2.2 Sample size estimation, recruitment, ethical considerations and data Collection Process

Sample size was calculated using the single proportion formula for the present study (Daniel formula, 1995): n = Z^2^ P (1-P)/d^2^ which gave 360 participants. By adding 10% we found a total of 396 patients with mental illness who were recruited from outpatient clinics of Kampala International University-Teaching Hospital and Jinja Regional Referral Hospital. Patients were chosen using simple random sampling method.

Patients who came for psychiatric review at outpatient clinics and fulfilling the inclusion criteria were approached and explained the purpose of the study. Approval to carry out the study was sought and obtained from Research Ethics committee of Bishop Stuart University (the research number is: BSU-REC-2023-99) and it has been conducted according to the principles expressed in the declaration of Helsinki. Before data collection, written informed consent from participants was obtained after fully explaining the details of the study to them in English and local Languages. Before asking consent from the patients, we first assessed their mental capacity using the Functional test of capacity. Participant were not forced to enroll themselves if they don’t want to. Participants were free to withdraw from the study any time he/she wishes without coercion or compromise of care they are entitled to. As patients with mental illness are among the vulnerable population, a written informed consent was also obtained from the next of kin and/ legally authorized representative for study participation.

Those who consented to participate in the study were given a questionnaire with sociodemographic informations, clinical characteristics, Medication Adherence Rating Scale (MARS) and the Ten Item Personality Inventory (TIPI) scale. Identification of participants was by means of codes.

The sociodemographic informations included age, gender, residence, level of education, marital status, employment status and family support. The questionnaire included also the diagnosis of the patient (confirmed by clinical interview using DSM-V-TR diagnostic criteria), associated comorbidities, duration of illness and duration of continuous therapy, number of tablets taken by the patient per day, the frequency as well as side effects of medicines.

Patients’ medication adherence was assessed using Medication Adherence Rating Scale (MARS). The MARS comprised of 10 items with response of yes or no and a total score of 10. A total score then revealed the level of adherence, scoring less than 6 was considered as non-adherence while 6 and above was considered as adherence (17).

To assess the personality traits, we used the Ten-Item Personality Inventory, which is a short version of Big Five Inventory. It consists of ten pairs of adjectives generated using a 7-point likert scale, with 7 (strongly agree) being the highest and 1 being the lowest (strongly disagree) (18). TIPI scoring (‘‘R’’ denotes reverse-scored items): Extraversion (1, 6R); Agreeableness (2R, 7); Conscientiousness (3, 8R); Neuroticism (4, 9R); Openness to Experiences (5, 10R). The score of the personality trait is the average of the 2 items. If the score of that personality trait is above the mean, the person was scored high on that trait. The questionnaire and the tools were pre-tested before being used in the study.

### 2.3 Data Processing and analysis

After being prepared and encoded, data were entered in Microsoft Excel version 2016 before being exported to Stata 15 for analysis. Proportions, percentages, and frequencies were calculated and presented in a table. Both bivariate and multivariate logistic regression analysis were used to assess personality traits and others factors associated with psychotropic medication adherence. For the multivariate analysis, variables that had a p-value below 0.2 at bivariate analysis were taken into consideration. The odds ratio (ORs) for the association were also presented, along with the appropriate 95% CI and p-values. Factors with a p-value was less than 0.05 were significantly associated with non-adherence. The personality trait for which p-value was less than 0.05 after controlling for the other possible factors was considered as significantly associated with non-adherence to psychiatric drugs.

## 3. Results

During the study period, to fulfil the expected sample size of 396 participants, we approached 410 patients with mental illness during the study period but only 397 were fulfilling the inclusion criteria among patients who came for review. One did not consent to participate in the study. The 396 patients that consented to participate in the study had the questionnaire filled and adherence assessed.

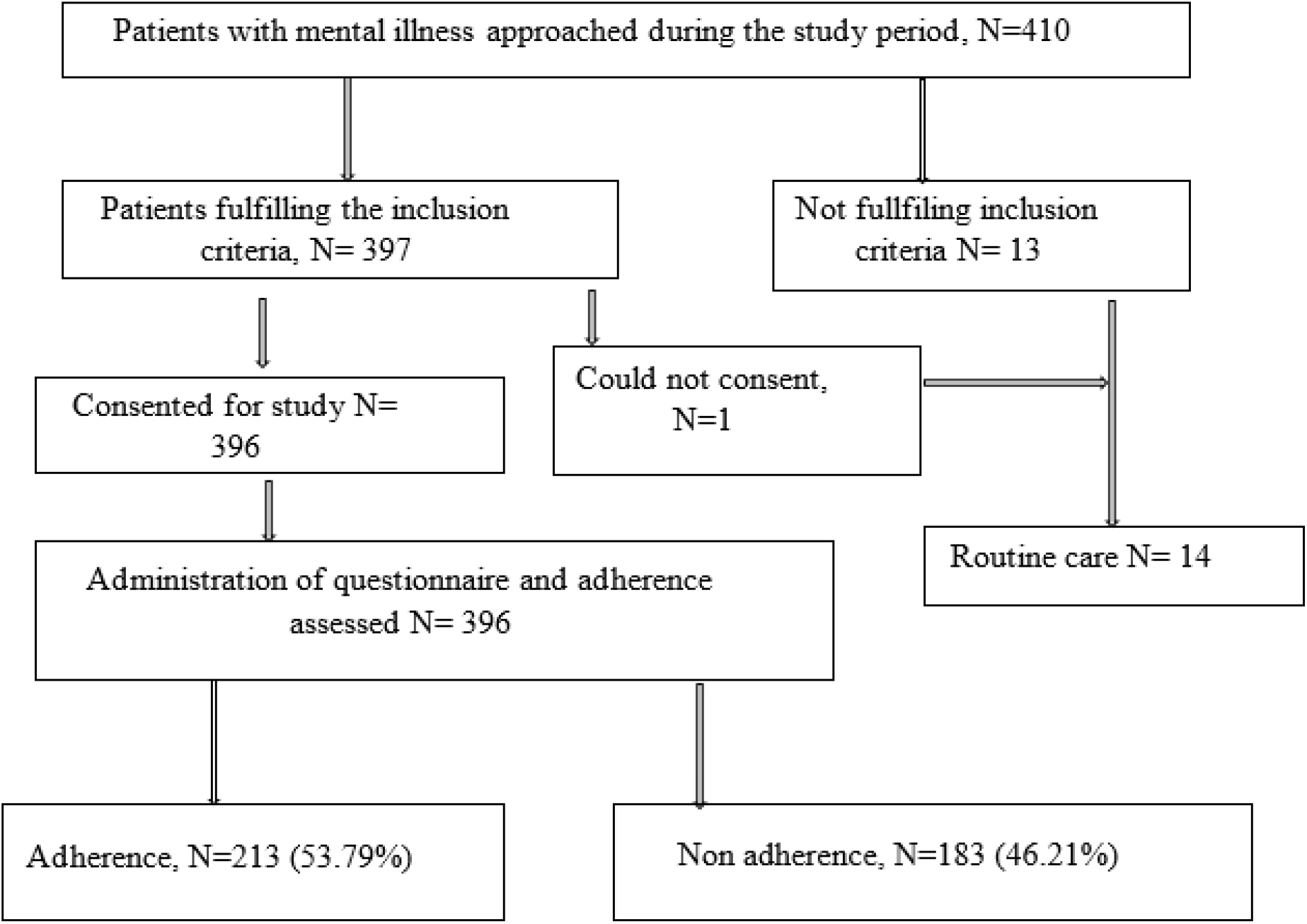

### 3.1. Sociodemographic and clinical characteristics of study participants

Males made up 234 (59.1%) of the study participants, with the majority falling into the 18–30 (42.7%) and 31–45 (41.2%) age categories. Majority were from pastoral areas 294(74.2%) and had the mental illness for 1-5 years (71.2%). The other baseline characteristics are shown in table 1(A) and 1(B) below

**Table 1(A):**
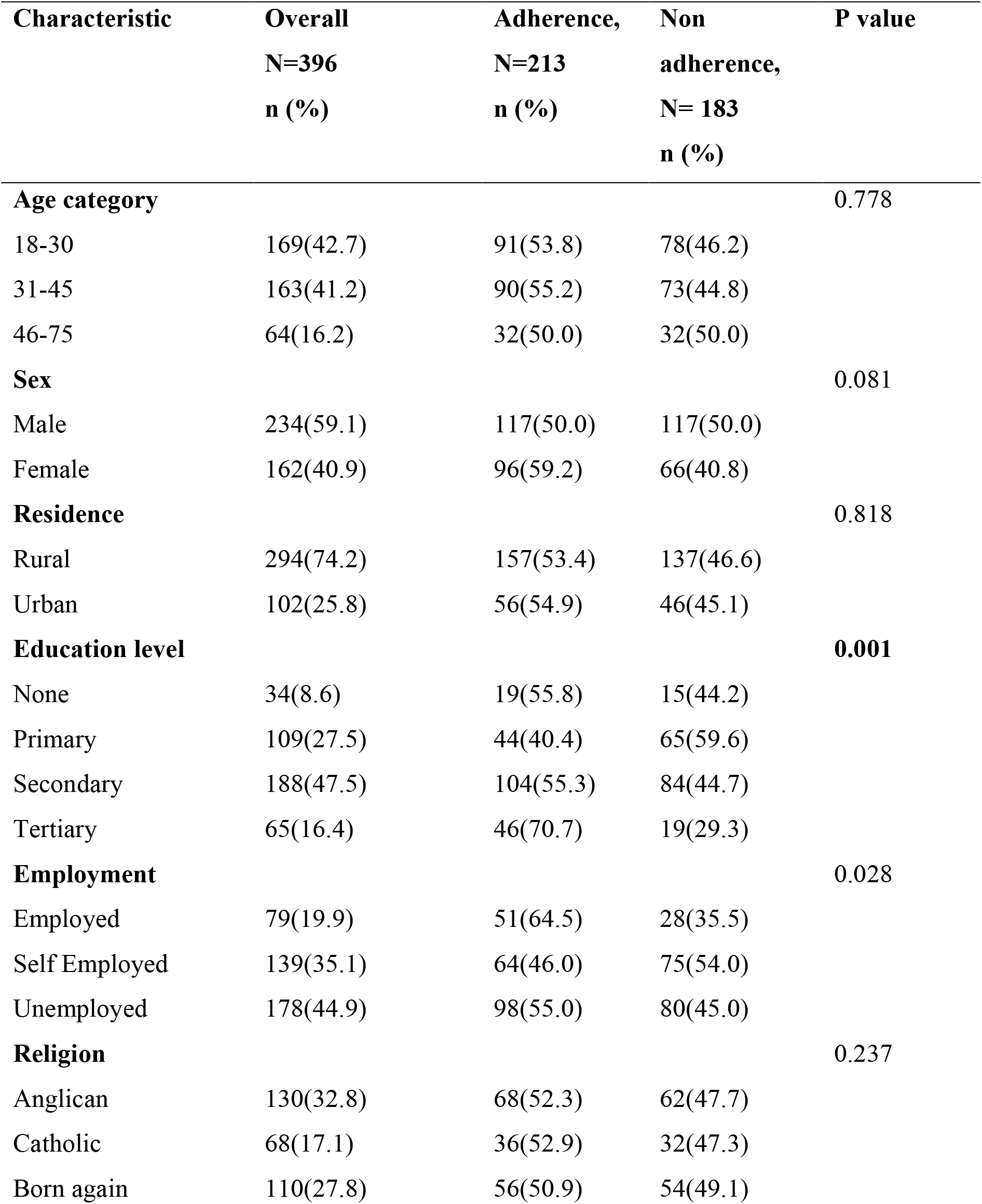
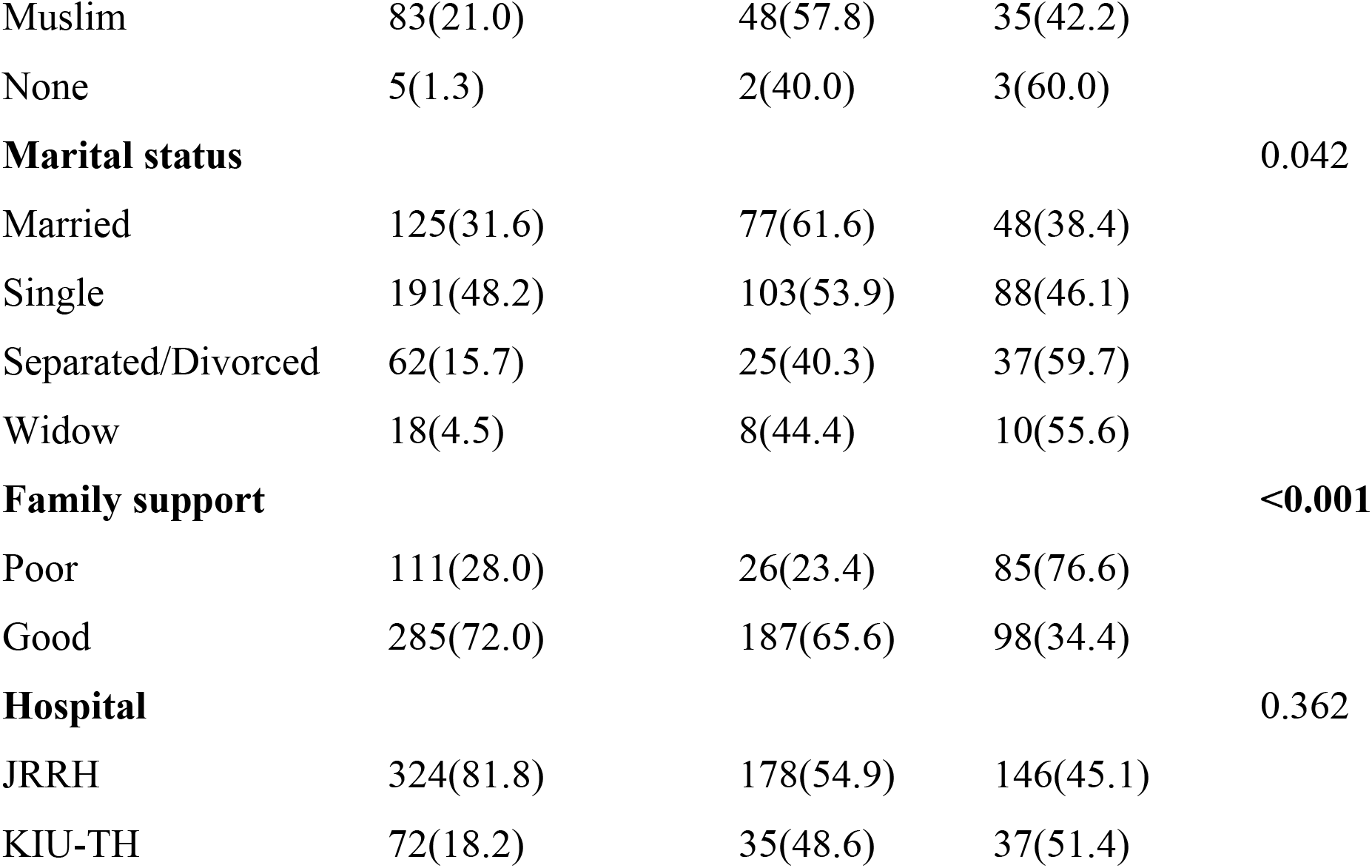
Sociodemographic characteristics of study participants.

**Table 1(B).**
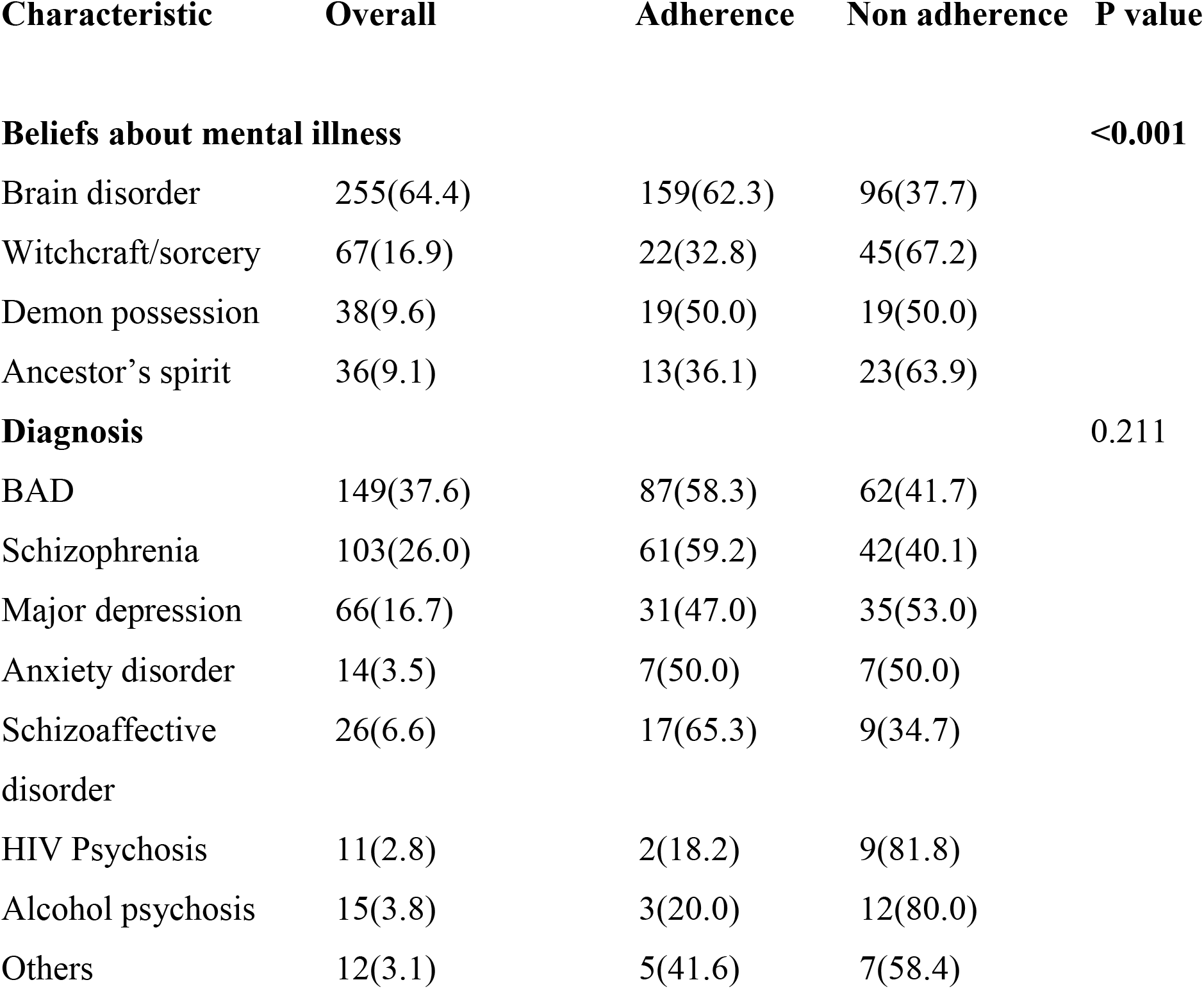
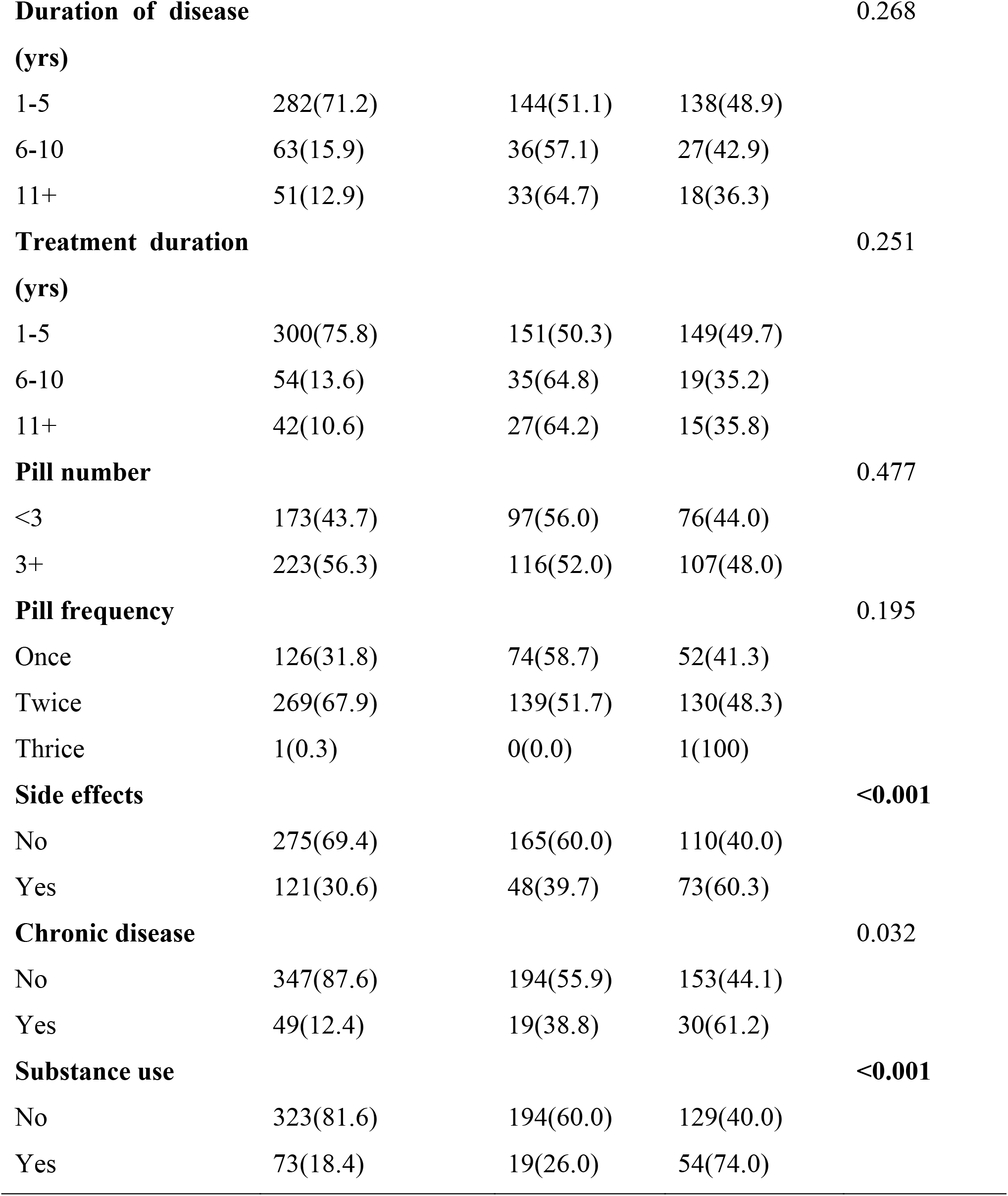
Clinical characteristics of study participants.

### 3.2. Prevalence of psychotropic medication adherence among people with mental illness at KIU-TH and JRRH

In this study, 213 patients were adherent while 183 were non adherent to psychotropic medication translating to 53.79% (95%CI 49.2-58.6) for adherence and 46.21(95%CI 41.4-50.8) for non-adherence respectively.

### 3.3. Sociodemographic and clinical factors associated with psychotropic medication adherence among people with mental illness at KIU-TH and JRRH

At bivariate level of analysis, the variables that had a p value less than 0.2, and therefore qualified for multivariate analysis were: sex, education level, employment status, marital status, family support, beliefs about mental illness, pill frequency, side effects, chronic disease and substance abuse.

At multivariate level of analysis, the variables that were independently associated with psychotropic medication non-adherence were poor family support, belief that mental illness is caused by witchcraft/sorcery, having side effects and substance use. The rest of the details of multivariate analysis are shown in table 2 below.

**Table 2:** Multivariate analysis of sociodemographic and clinical factors associated with psychotropic medication non-adherence among people with mental illness at KIU-TH and JRRH.

| Characteristic | Bivariate analysis |  |  |  | Multivariate analysis | P value |
| --- | --- | --- | --- | --- | --- | --- |
|  | cOR | 95%CI | P value | aOR | 95%CI |  |
| **Sex** |  |  |  |  |  |  |
| Male | 1.455 | 0.970-2.180 | 0.070 | 1.069 | 0.643-1.778 | 0.796 |
| Female | Ref |  |  |  |  |  |
| **Education level** |  |  |  |  |  |  |
| None | 1.911 | 0.807-4.528 | 0.141 | 0.869 | 0.281-2.691 | 0.808 |
| Primary | 3.577 | 1.853-6.902 | <0.001 | 1.864 | 0.790-4.396 | 0.155 |
| Secondary | 1.955 | 1.066-3.588 | 0.030 | 1.296 | 0.600-2.801 | 0.509 |
| Tertiary | Ref |  |  |  |  |  |
| **Employment** |  |  |  |  |  |  |
| Employed | Ref |  |  |  |  |  |
| Self Employed | 2.134 | 1.208-3.771 | 0.009 | 1.412 | 0.684-2.913 | 0.350 |
| Unemployed | 1.487 | 0.860-2.571 | 0.156 | 0.942 | 0.468-1.893 | 0.866 |
| **Marital status** |  |  |  |  |  |  |
| Married | Ref |  |  |  |  |  |
| Single | 1.371 | 0.866-2.169 | 0.179 | 0.712 | 0.401-1.263 | 0.245 |
| Separated | 2.374 | 1.274-4.424 | 0.006 | 0.721 | 0.315-1.648 | 0.438 |
| Widow | 2.005 | 0.740-5.435 | 0.171 | 0.568 | 0.157-2.064 | 0.390 |
| **Family support** |  |  |  |  |  |  |
| **Poor** | **6.238** | **3.774-10.313** | **<0.001** | **6.915** | **3.679-12.998** | **<0.001** |
| Good | Ref |  |  |  |  |  |
| **Beliefs about mental illness** |  |  |  |  |  |  |
| Brain disorder | Ref |  |  |  |  |  |
| **Witchcraft/Sorcery** | **3.388** | **1.917-5.987** | **<0.001** | **2.959** | **1.488-5.884** | **0.002** |
| Demon possession | 1.656 | 0.835-3.284 | 0.149 | 1.306 | 0.591-2.887 | 0.510 |
| Ancestor's spirit | 2.930 | 1.418-6.055 | 0.004 | 2.088 | 0.871-5.009 | 0.099 |
| **Pill frequency** |  |  |  |  |  |  |
| Once | Ref |  |  |  |  |  |
| Twice | 1.331 | 0.868-2.041 | 0.190 | 0.885 | 0.514-1.523 | 0.658 |
| **Side effects** |  |  |  |  |  |  |
| No | Ref |  |  |  |  |  |
| **Yes** | **2.281** | **1.474-3.531** | **<0.001** | **2.257** | **1.326-3.843** | **0.003** |
| **Chronic disease** |  |  |  |  |  |  |
| No | Ref |  |  |  |  |  |
| Yes | 2.002 | 1.085-3.694 | 0.026 | 1.874 | 0.895-3.920 | 0.096 |
| **Substance use** |  |  |  |  |  |  |
| No | Ref |  |  |  |  |  |
| **Yes** | **4.274** | **2.421-7.545** | **<0.001** | **4.174** | **2.121-8.214** | **<0.001** |

*aOR= Adjusted odds ratio, Ref= Reference category, CI= Confidence interval*

### 3.4. Personality traits associated with psychotropic medication adherence among people with mental illness

At bivariate analysis, only neuroticism and agreeableness were the two personality with p value less than 0.2 and qualified for multivariate analysis (Table 3).

**Table 3:**
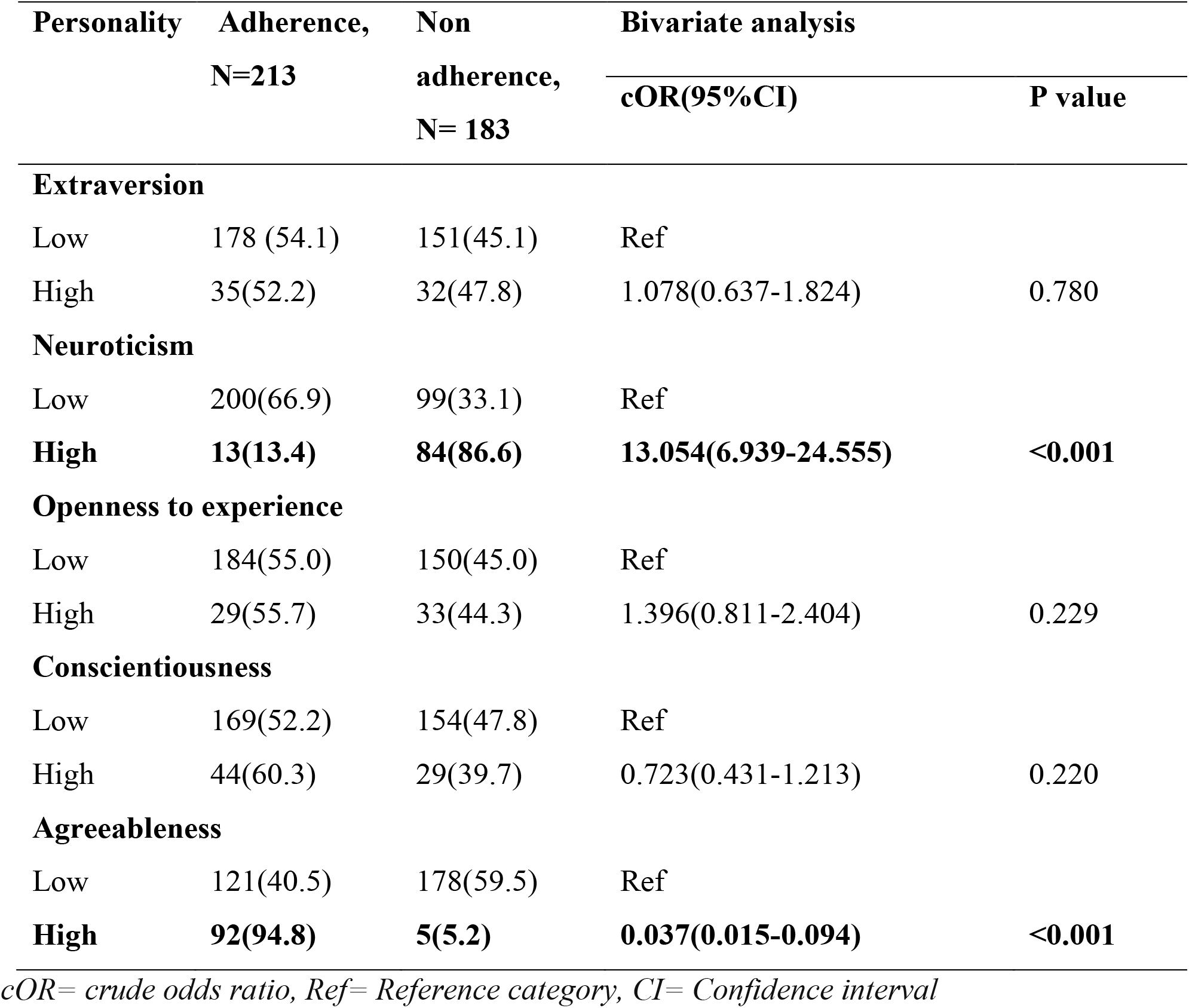
Bivariate analysis of traits associated with psychotropic medication non-adherence among people with mental illness at KIU-TH and JRRH.

At multivariate analysis after controlling for potential confounders which included absence of family support, belief that mental illness is caused by witchcraft/sorcery, having side effects and substance use, agreeableness and neuroticism personality traits remained significantly associated with non-adherence. A participant with personality trait neuroticism was 7.424 times more likely to be non-adherent (aOR=7.424, CI=3.890-14.168, P<0.001). A participant with personality trait agreeableness was 0.062 times less likely to be non-adherent (aOR=0.062, CI=0.024-0.160, P<0.001). This means that a participant with personality trait agreeableness was 16.12 (1/0.062) times more likely to be adherent (Table 4).

**Table 4:** Multivariate analysis of personality traits associated with psychotropic medication non-adherence among people with mental illness at KIU-TH and JRRH (controlling for potential confounders). aOR= Adjusted odds ratio, Ref= Reference category, CI= Confidence interval.

| Characteristic | Bivariate analysis |  |  | Multivariate analysis |  |  |
| --- | --- | --- | --- | --- | --- | --- |
|  | cOR | 95%CI | P value | aOR | 95%CI | P value |
| **Neuroticism** |  |  |  |  |  |  |
| Low | Ref |  |  |  |  |  |
| **High** | **13.054** | **6.939-24.555** | **<0.001** | **7.424** | **3.890-14.168** | **<0.001** |
| **Agreeableness** |  |  |  |  |  |  |
| Low | Ref |  |  |  |  |  |
| **High** | **0.037** | **0.015-0.094** | **<0.001** | **0.062** | **0.024-0.160** | **<0.001** |
| **Family support** |  |  |  |  |  |  |
| No | **6.238** | **3.774-10.313** | **<0.001** | **6.915** | **3.679-12.998** | **<0.001** |
| Yes | Ref |  |  |  |  |  |
| **Beliefs about mental illness** |  |  |  |  |  |  |
| Brain disorder | Ref |  |  |  |  |  |
| Witchcraft/Sorcery | **3.388** | **1.917-5.987** | **<0.001** | **2.867** | **1.385-5.682** | **0.002** |
| Demon possession | 1.656 | 0.835-3.284 | 0.169 | 1.348 | 0.598-2.896 | 0.516 |
| Ancestor's spirit | 2.930 | 1.418-6.055 | 0.004 | 2.064 | 0.865-5.018 | 0.096 |
| **Side effects** |  |  |  |  |  |  |
| No | Ref |  |  |  |  |  |
| Yes | **2.281** | **1.474-3.531** | **<0.001** | **2.242** | **1.322-3.838** | **0.001** |
| **Substance use** |  |  |  |  |  |  |
| No | Ref |  |  |  |  |  |
| Yes | **4.274** | **2.421-7.545** | **<0.001** | **4.146** | **2.114-8.204** | **<0.001** |

## 4. Discussion

In this hospital-based cross-sectional study we aimed to determine the prevalence of psychotropic medication non-adherence and associated personality traits (and other factors) among people with mental illness

In our study, the prevalence of psychotropic medication non-adherence was 46.21(95%CI 41.4-50.8). The high prevalence of medication non-adherence found in our study can also be explained by the false beliefs about mental illness and the use of alternative medicine in Uganda, among others factors.

Our prevalence was in in range of previous studies done in Nigeria and Malawi. In Nigeria study done by Adewuya has shown that 48% of the participants were non-adherent to medications (19) and Mekani in Malawi has found that 46.2% of the study participants were adherent to psychotropic medication (20). However, the prevalence of non-adherence in our study was higher than the prevalence of medication non-adherence found by Gudeta (37.7%) and Mekuriwa (39.3%) in Ethiopia as well as 36.4% found by Ibrahim in Nigeria (22;23;24). The prevalence was lower than 61.8% found by Nega et al. in Ethiopia and 58.7% found in India by Nagesh et al. (25;26).

This difference could result from the difference in study settings and the tools used to assess medication adherence and personality traits.

### Personality traits associated with psychotropic medication non-adherence

In our study, the two personality traits that were significantly associated with drug non-adherence after controlling for the significant sociodemographic and clinical factors were Neuroticism and Agreeableness.

In our study participant with neuroticism traits were more likely to be non-adherent to psychotropic medications. Persons with predominant neuroticism personality traits are likely to have instability in emotions, self-doubt, irritability, anxiety, and depression. They experience generally negative emotions which can lead to poor adherence to medications. Our results were in line with other studies done in different parts of the world (21). However, a study done in Italy using the short version of Big Five Inventory (BFI-10) to assess personality traits has found that conscientiousness was also associated with non-adherence to medication (21). The difference can be explained by the small simple size (13 participants) and the longitudinal nature of the study.

Agreeableness was also significantly associated with psychotropic medication non-adherence. A participant with predominant agreeableness traits was less likely to be non-adherent to medications. The patients with predominant agreeableness personality traits tend to agree and follow other’s opinions which can help to adhere to instructions given by health workers and as result the increase in the level of adherence to medications.

This is similar to the results of a study done in Sweden who found that agreeableness influences positively the adherence behavior (21) and a systematic review done in India, studies showed that Agreeableness was among the personality traits negatively associated with non-adherence for patients with chronic conditions (21).

However, in USA, the most personality traits associated with non-adherence among participants with bipolar affective disorder were extraversion, openness and conscientiousness (21). This difference was due to the study population (our study was done in all patients with different mental disorders not only in patients with bipolar affective disorder) and also, he used the revised Five Factor Inventory (Neo-FFI) with 60 items. In U.K only conscientiousness was the personality trait influencing non-adherence behavior (21).

This difference may be explained by the different versions of the Big Five Inventory used to assess personality traits, the methodology used in the study (longitudinal and multicenter) but also the sample size (1062 patients).

Other factors associated with psychotropic medication non-adherence were significantly associated with medication non-adherence were poor family support, belief that mental illness is caused by witchcraft/sorcery, presence of side effects and substance use.

Poor family support had negative influence on medication adherence. In Uganda where there is lack of social protection agencies, families are the principal sources of support and many families in low-income countries have social and financial difficulties (22). Family support improve medication adherence by taking care of the patient, reminding him to take medicine, providing basics needs and money for transport to hospital or for medicine. The false beliefs about causes of mental illness influenced psychotropic medication non-adherence. The beliefs about the causes of mental illness influence the choice to adhere to modern treatment (23) which lead to poor compliance to biological treatment. The false beliefs about the causes of mental illness lead to poor insight about illness and poor insight leads to psychotropic medication non-adherence (24). The presence of side effects of medication was significantly associated with medication non-adherence in our study. Individuals who experienced side effects were more likely to be non-adherent compared to those without side effects. Patients who experience severe side effects can intentionally stop medication to relieve the side effects. Substance use was also significantly associated with psychotropic medication non-adherence in our study. Patients who used psychoactive substances were more likely to be non-adherent compared to others. Psychoactive substance impair cognition and rationale thinking and are also associated with increased adverse effects and interactions with psychotropic medications (25) which can increase non-adherence to psychotropic medications.

### Strengths of the study

- The study was done in public and private referral hospitals; the findings may apply to both public and private institutions.
- This is the first study exploring the association between personality traits and psychotropic medication non-adherence in Uganda and in the East African region.

### Limitations of the study

- The study was cross-sectional which measures data at a single point in time
- The study was done in only two hospitals chosen by convenience, so the results of the study cannot be generalized for the country.
- The tools used were self-reported which can decrease the accuracy of the responses.

### Areas of further research

Longitudinal studies on several sites of the country are needed to generalize the results for the country.

## Conclusion

Medication non-adherence was high. Non-adherent patients were more likely to have predominant neuroticism personality traits. Medication non-adherence was shown to be less common in individuals with agreeableness personality traits. Other factors associated with medication non-adherence were poor social support, witchcraft beliefs, side effects and substance abuse. Reinforced psycho-education when giving medication to patients with high risk of non-adherence and community based interventions to increase awareness about mental illnesses could help to decrease non-adherence to psychotropic medication non-adherence.

## Data Availability

All relevant data are within the manuscript and its Supporting Information files.

## List of abbreviations

JRRH: Jinja Regional Referral Hospital
KIU-TH: Kampala International University Teaching hospital
MARS: Medication Adherence Rating Scale
TIPI: Ten Items Personality Inventory

## Declarations

### Ethical approval and consent to participate

Approval to carry out the study was sought and obtained from Research Ethics committee of Bishop Stuart University (BSU-REC-2023-99).Written informed consent from participants was obtained after fully explaining the details of the study to them in English and local Languages and assessing capacity to consent using the functional test for capacity.Participant were not forced to enroll themselves if they don’t want to. Participants were free to withdraw from the study any time he/she wishes without coercion or compromise of care they are entitled to. As patients with mental illness are among the vulnerable population, a written informed consent was also obtained from the next of kin and/ legally authorized representative for study participation.

### Consent for publication

Not applicable.

### Authors’ contributions

- Emmanuel Niyokwizera, principal investigator has conceived the study, elaborated study design and executed it from the beginning of the study to the submission of the manuscript.
- Kirabira Joseph and Nnaemka Chukwudum Abamara have supervised the study from the conception till submission of the manuscript.
- Niyubahwe Raissa and David Nitunga have helped in data collection and edition of the article
- Joshua Muhumuza has helped in data analysis.

## Acknowledgements

The research assistants Waiswa Daniel (psychiatric clinical officer JRRH) participated in data collection. We appreciate also all the patients who accepted to participate in the study

## Availability of data and materials

The data and materials used in this study are all available within the manuscript and supporting files.

## References

1. Trautmann S, Wittchen H. The economic costs of mental disorders:Do our societies react appropriately to the burden of mental disorders ? EMBO Rep. 2016;17(9):1245– 9.

2. Charlson FJ, Diminic S, Lund C, Degenhardt L, Whiteford HA. Mental and substance use disorders in Sub-Saharan Africa: Predictions of epidemiological changes and mental health workforce requirements for the next 40 years. PLoS One. 2014;9(10):1– 11.

3. WHO. Depression and Other Common Mental Disorders Global Health Estimates. Glob Heal Estim Geneva. 2017;2017(pp.):1–24.

4. Nelson Opio J, Tufanaru C, Aromataris E. Prevalence of mental disorders in Uganda: A systematic review protocol. JBI Database Syst Rev Implement Reports. 1. 2018;16(8):1613–20.

5. Abbo C. Profiles and outcome of traditional healing practices for severe mental illnesses in two districts of Eastern Uganda. Glob Health Action. 2011;4(7117):1–15.

6. Semahegn, A., Torpey, K., Manu, A., Assefa N; Tesfay. Psychotropic medication non-adherence and its associated factors among patients with major psychiatric disorders: A systematic review and meta-analysis. Syst Rev. 2020;9(1):1–18.

7. Gebeyehu DA, Mulat H, Bekana L, Asemamaw NT, Birarra MK, Takele WW, et al. Psychotropic medication non-adherence among patients with severe mental disorder attending at Bahir Dar Felege Hiwote Referral hospital, north west Ethiopia, 2017. BMC Res Notes. 2019;12(1):2–7.

8. Chukwujekwu CD, Adesokun OK. Prevalence of Medication Non-Adherence among Psychiatric Patients in a Tertiary Hospital in Nigeria. J Biosci Med. 2017;05(04):1–10.

9. Hugtenburg JG, Timmers L, Elders PJ, Vervloet M, Dijk L van. Definitions, variants, and causes of nonadherence with medication : a challenge for tailored interventions. Dovepress J patient Prefer adherence. 2013;7:675–82.

10. Rababa’h S, Yousef F. Predictors of Non-adherence in Patients Taking Psychotropic Medication and Suggestions to Improve. J Heal Med Nurs. 2017;38(2):73–84.

11. Nsereko ND. The evolution of mental health understanding and practice in Uganda. Int J Emerg Ment Health. 2017;19(1):1–6.

12. Adeniran A, Akinyinka M, Wright KO, Bakare OQ, Goodman OO, Kuyinu YA, et al. Personality Traits, Medication Beliefs & Adherence to Medication among Diabetic Patients Attending the Diabetic Clinic in a Teaching Hospital in Southwest Nigeria. J Diabetes Mellit. 2015;05(04):319–29.

13. Atherton OE, Willroth EC, Schwaba T, Goktan AJ, Graham EK, Condon DM, et al. Personality predictors of emergency department post-discharge outcomes. Personal Sci. 2021;2:1–20.

14. Ferretti F, Laurenzi PF, Luca A De. Changes in Adherence to Oral Medications of People with Mental Illness During the Pandemic: Neuroticism and Conscientiousness Personality Traits Moderate the Effect of Medication Beliefs. Rresearch Sq. 2022;10(59):1–13.

15. Kohli R. A Systematic Review to Evaluate the Association Between Medication Adherence And Personality Traits. Value Heal. 2017;20(9):A686.

16. Okasha TA, Radwan DN, Elkholy H, Hendawy HMFM, Shourab1 EMME, Teama1 RRA, et al. Psycho-demographic and clinical predictors of medication adherence in patients with bipolar I disorder in a university hospital in Egypt. South African J psychiatry. 2020;26(0):1–9.

17. Iseselo MK, Ambikile JS. Medication challenges for patients with severe mental illness: Experience and views of patients, caregivers and mental health care workers in Dar es Salaam, Tanzania. Int J Ment Health Syst. 2017;11(1):1–12.

18. Thompson K, Kulkarni J, Sergejew AA. Reliability and validity of a new Medication Adherence Rating Scale (MARS) for the psychoses Reliability and validity of a new Medication Adherence Rating Scale (MARS) for the psychoses k. Schizophr Res. 2000;42:241–247.

19. Gosling SD, Rentfrow PJ, Jr WBS. A very brief measure of the Big-Five personality domains q. J Res Pers. 2003;37:504–28.

20. Adewuya AO, Owoeye OA, Erinfolami AR, Ayodele O. Prevalence and correlates of poor medication adherence amongst psychiatric outpatients in southwestern Nigeria. Gen Hosp Psychiatry. 2010;32(1)(116):95–6.

21. Mekani P. Clinic appointments and medication adherence among patients accessing mental health services in Central Malawi : A cross-sectional analytical study. Resarch Sq. 2023;1(2):1–17.

22. Gudeta DB, Leta K, Alemu B, Kandula UR. Medication adherence and associated factors among psychiatry patients at Asella Referral and Teaching Hospital in Oromia, Ethiopia: Institution based cross sectional study. PLoS One. 2023;18(4):e0283829.

23. Mekuriaw B, Tessema W, Agenagnew L, Dawud B, Abdisa E, Tesfaye M. Medication Non-adherence and Use of Traditional Treatment Among Adult Psychiatric Patients in Jimma Town Treated at Jimma University Teaching. J Psychiatry. 2016;21(1):1–7.

24. Ibrahim AW, Yahya S, Pindar SK, Wakil MA, Garkuwa A SS. Prevalence and predictors of sub-optimal medication adherence among patients with severe mental illnesses in a tertiary psychiatric facility in Maiduguri, North-eastern Nigeria. ssPanafrica Med J. 2015;21(39):1–11.

25. Nega M, Demissie M, Semahegn A, Tesfaye G. Psychotropic Medication Non-Adherence among Psychiatric Patients at Two Hospitals in Eastern Ethiopia. East African J Heal Biomed Sci. 2018;2(November):45–54.

26. Nagesh HN, Kishore MS, Raveesh BN. Assessment of adherence to psychotropic medications in a psychiatric unit of district hospital. Natl J Physiol Pharm Pharmacol. 2016;6(6):581–5.

27. Antiri EO, Ocansey S, Ntodie M, Abokyi S, Abraham CH. Associations Between Personality Traits and Adherence to Treatment in Patients with Primary Open Angle Glaucoma in an African Population. Resarch Sq. 2021;10(75):1–13.

28. Axelsson M, Brink E, Lundgren J, Lötvall J. The influence of personality traits on reported adherence to medication in individuals with chronic disease: An Epidemiological study in West Sweden. PLoS One. 2011;6(3):1–7.

29. Sachs GS, Peters AT, Sylvia L, Grunze H. Polypharmacy and bipolar disorder: What’s personality got to do with it? Int J Neuropsychopharmacol. 2014;17(7):1053–61.

30. Kirchner S. Medication Adherence in a Cross-Diagnostic Sample of Patients From the Affective-to-Psychotic Spectrum: Results From the PsyCourse Study. Front Psychiatry. 2022;12(01):1–10.

